# Rapid detection of non-rifampicin drug resistant tuberculosis using Xpert MTB/XDR testing: Findings from a multisite drug-resistant tuberculosis case detection surge in Zambia

**DOI:** 10.1101/2025.11.11.25340045

**Authors:** Phallon B. Mwaba, Marshal Sikandangwa, Robertson Chibumbya, Innocent Mwaba, Minyoi M. Maimbolwa, Chitalu Chanda, Muhammad Mputu, Rhehab Chimzizi, Kevin M. Zimba, Nancy Kasese-Chanda, Angel Mubanga, Monde Muyoyeta, Mary Kagujje

## Abstract

**Background:** Zambia continues to face a high burden of tuberculosis with rising drug resistance, yet diagnosis of non-rifampicin resistance remains limited due to reliance on rifampicin as a proxy for multidrug-resistant tuberculosis. The Xpert MTB/XDR assay offers rapid detection of resistance to isoniazid, fluoroquinolone, Ethionamide and aminoglycosides, addressing a major diagnostic gap. This study assessed the feasibility and utility of reflex MTB/XDR testing using data from a drug-resistant tuberculosis active case finding surge.

**Methods:** We conducted an analysis of programmatic data from a four-week active case finding surge in May 2025, implemented across 39 high-TB burden facilities in five provinces of Zambia. We evaluated bacteriologically confirmed (Xpert MTB/RIF Ultra) tuberculosis patients using MTB/XDR regardless of Rifampicin resistance. We analyzed patient demographics, characteristics, and drug-resistance patterns descriptively.

**Results:** Of 509 TB patients identified, 70.9%(n=361) were bacteriologically confirmed by Xpert MTB/RIF Ultra among whom 92.8%(n=335) were eligible for reflex Xpert MTB/XDR testing. Of the eligible, 89.3% (n=299) underwent reflex MTB/XDR testing. Results were successfully retrieved for 282 (94.3%). Overall, 8.5%(n=24) of patients with XDR results had a form of drug-resistant tuberculosis. Mono-resistance to rifampicin (3.5%, n=10) or isoniazid (3.2%, n=9) was more prevalent than multi-drug-resistant tuberculosis (1.1%, n=3), while only 0.4% (n=1) had Pre-XDR TB. Non-rifampicin resistance, including isoniazid mono-resistance and isoniazid combined with fluoroquinolone or ethionamide resistance, were detected in 3.2% (n=9) of all patients with XDR TB results (37.5% of all DR-TB cases).

**Conclusion:** Reflex Xpert MTB/XDR testing enabled rapid identification of non-rifampicin-resistant TB cases that would have been missed by rifampicin-centric diagnostics. Strengthening the integration of MTB/XDR into routine diagnostic algorithms can improve early drug-resistant tuberculosis detection, guide individualized treatment, and prevent amplification of resistance in high-burden settings. National scale-up, accompanied by laboratory strengthening, capacity building, and robust data systems, are essential to optimizing its use and advancing drug-resistant tuberculosis elimination.

## Introduction

Zambia, one of the 30 high tuberculosis (TB) burden countries globally, continues to shoulder a significant TB disease burden with a high incidence of drug-resistance (DR). In 2023, the World Health Organisation (WHO) estimated Zambia’s TB incidence at 283 per 100,000 population with multi-drug or rifampicin-resistant (MDR/RR) TB constituting 1.3% of new and 9.1% of retreatment cases (1). Despite improvements in TB elimination efforts such as increased drug-resistant (DR) TB surveillance through expanded GeneXpert coverage countrywide, Zambia continues to face challenges in diagnosing DR TB potentially contributing to a steady decline DR TB notifications since 2018 (1). In 2023, an estimated 1673 people developed MDR/RR TB, yet only 403 (23.7%) were confirmed in the laboratory (1) suggesting a huge number of missing DR TB patients.

Like in other resource limited settings, the under-diagnosis of DR TB is attributable to limited access to advanced molecular diagnostics as well as weaknesses in sample transportation and laboratory network to facilitate culture and drug susceptibility testing in reference laboratories (2). While the use of the GeneXpert MTB/RIF Ultra has improved rifampicin (RIF) resistance detection, it fails to detect other first-line resistance patterns including isoniazid (INH) and second-line DR such as ethionamide (ETH), fluoroquinolones (FQs) and aminoglycosides such as kanamycin and amikacin (3). This gap poses a significant risk: individuals with non-RIF resistance may be erroneously treated with first-line regimens, potentially leading to treatment failure, amplification of resistance, mortality and continued community transmission (4,5). Therefore, rapid and comprehensive DR TB diagnostics are essential in high-burden settings.

The recently introduced WHO-recommended Xpert MTB/XDR assay builds upon the established Xpert MTB/RIF Ultra platforms to provide rapid, cartridge-based detection of resistance to INH, FQs, ETH and aminoglycosides (amikacin, kanamycin and capreomycin) within hours (6,7). This tool has shown high sensitivity and specificity across diverse settings and is recommended for use in patients at risk of second-line DR TB or those with prior treatment history (8–10) and as a reflex test for all bacteriologically confirmed TB (3).

Although Zambia introduced Xpert MTB/XDR testing in programmatic setting in 2024, its uptake remains limited, and non-RIF resistance patterns are still poorly characterized. In response, the United States Government funded Tuberculosis Local Organizations Network (TBLON) project supported the Ministry of Health (MoH) to conduct a targeted DR-TB active case finding (ACF) surge in May 2025. This was conducted across project-supported high TB burden sites that incorporated Reflex Xpert MTB/XDR testing for eligible bacteriologically confirmed TB patients. We report programmatic data from this intervention to quantify the prevalence of non-rifampicin resistance among bacteriologically confirmed TB patients regardless of the RIF resistance status.

## Materials and Methods

### Intervention Design and Setting

This was an analysis of programme data from a DR TB ACF surge conducted between 5^th^ May and 31^st^ May 2025. The surge was a targeted, multi-site intervention conducted by Zambia’s National TB and Leprosy Programme (NTLP) with support from the TBLON project. The surge was implemented in 39 high-TB health facilities in five high TB burden provinces in Zambia: Lusaka, Southern, Copperbelt, Northern and Central Provinces.

Provinces and facilities were selected based on their DR-TB burden: provinces with ≥25 DR TB notifications and health facilities with at least five (5) DR TB notifications in 2024 were selected to participate in the surge.

The participating health facilities routinely conduct DR TB surveillance activities through clinical screening and sample evaluation using GeneXpert MTB/RIF Ultra. Although reflex testing for all bacteriologically confirmed TB patients using Xpert MTB/XDR is required, it is not routinely available and performed.

### Intervention Procedures

The DR TB ACF surge had a preparatory and an implementation phase.

#### Preparatory Phase

After co-developing the concept note with the NTLP, pre-surge key components included planning, logistics management, development of digital monitoring tools, health care workers capacity building as well as community mobilization and demand generation.

To ensure commodity availability, we conducted facility-level assessments to determine baseline stocks of GeneXpert Ultra and Xpert MTB|XDR cartridges as well as TB drugs. Where necessary, MoH staff made emergency orders, and the project supported commodity transportation and distribution including redistributions. Furthermore, we developed a courier matrix to facilitate timely sample referral from participating facilities to Xpert MTB/XDR sites.

We developed a surge orientation package covering DR-TB definitions, case finding, integration of Xpert MTB|XDR testing, demand generation strategies and data entry procedures. Using this package, we capacity-built healthcare workers (HCWs) including clinicians, nurses, Community Health Workers (CBWs), and data staff through facility-based orientation sessions.

We implemented community mobilization and demand generation efforts throughout both the pre-surge and intra-surge periods. We developed a standardized community engagement script and applied it across interpersonal communication, media programs, and church visits to raise awareness of DR-TB, enhance service acceptability, and stimulate demand.

To facilitate adequate abstraction of information from registers and performance monitoring, the Strategic Information team developed an electronic data collection tool within REDCap version 14.8.2 for patient-level data management during the surge. The tool underwent iterative testing and revisions before rollout.

#### Implementation phase

Two categories of patients were targeted during this surge. The first category included TB patients diagnosed in the preceding two months using microscopy and urine LAM or who were smear positive at month 2 of treatment. During the surge, these patients were re-evaluated using GeneXpert MTB/RIF Ultra to determine their rifampicin resistance profile and eligibility for GeneXpert MTB/XDR. The second category included prospective bacteriologically confirmed TB patients, diagnosed using GeneXpert MTB/RIF Ultra. The Gene Xpert positive TB patients, from both categories, excluding those with trace call results, were further evaluated using GeneXpert MTB/XDR to rapidly detect resistance to INH, FQ, ETH and aminoglycosides. Where submitted samples were inadequate, fresh ones were collected for Xpert MTB/XDR testing. All diagnosed DR-TB patients received psychosocial counselling to promote treatment initiation and adherence and initiated on appropriate treatment regimens (e.g., BPaLM or RZE+Lfx for INH monoresistance) for DR-TB patients.

#### Monitoring, data management and analysis

Daily implementation was jointly monitored by TBLON and MoH teams through structured supervision visits that provided onsite support for data collection, quality assurance, and troubleshooting. Facility-level MoH staff routinely entered the data in the TB treatment and laboratory registers. De-identified data from the registers was digitized into REDCap v14.8.2. Twice-weekly virtual performance review meetings were held to analyze trends, address barriers, and guide adaptive management. Furthermore, we disseminated weekly data summaries to facilities to reinforce accountability and enable timely intervention where challenges were noted.

Data analysis was done using STATA v17.0 (StataCorp, USA) and conducted descriptive data analysis. We did not conduct any inferential statistical tests considering the small absolute numbers of DR-TB patients.

### Definition of Terms in the context of this study

i. Reflex Xpert MTB/XDR testing – A follow on GeneXpert MTB/XDR conducted on all bacteriologically confirmed TB patients on residual specimens previously processed for Xpert MTB/RIF Ultra test or a new one if the initial sample was inadequate (11).
ii. MDR - Resistance to both isoniazid and rifampicin with or without resistance to other anti TB drugs (12)
iii. Mono-resistance– Resistance to only one first line drug (13).
iv. Pre-XDR TB – MTB that fulfils the definition of multidrug-resistant or rifampicin-resistant TB (MDR/RR-TB) and also resistant to any fluoroquinolone (14)
v. Poly-resistance - resistance to more than one first-line anti-TB drug, beside both rifampicin and isoniazid (12).

### Ethical Considerations

The TBLON project has ethical approval from the University of Zambia Biomedical Research Ethics Committee (REF, NO. 1773-2021) to use routinely collected program data. Waiver of informed consent was granted by UNZABREC as all analysis would be performed on de-identified routine program data.

## Results

We identified 509 TB patients (Fig 1) of which 160 (31.4%) were either previously diagnosed using smear microscopy/LAM or were smear positive at month 2 of treatment (category 1) and 349 (68.6%) were Xpert positive diagnosed during surge (Category 2). Among the category 1 patients, 148 (92.5%) were re-evaluated using GeneXpert during the surge with 11 (7.4%) testing positive for Mycobacterium tuberculosis (MTB). Among the 349 patients diagnosed with bacteriologically confirmed TB during the surge, 4.3% (n=15) had rifampicin resistant [RR] TB; 88.8% (n=310) RIF susceptible. Among patients with RR TB, 14 (93.3%) submitted samples for reflex XDR testing with 28.6% (n=4) detected with additional DR. Of the 310 RIF susceptible patients, 93.2% (n=289) submitted samples for reflex XDR testing to identify non-rifampicin resistance and results were successfully retrieved for 93.4% (n=271) patients (a total of 282 results from category 1 and category 2). Overall, 8.5% (n=24) of bacteriologically confirmed TB patients with accessible XDR results were found to have some form of DR TB (all forms).

**Figure 1.**
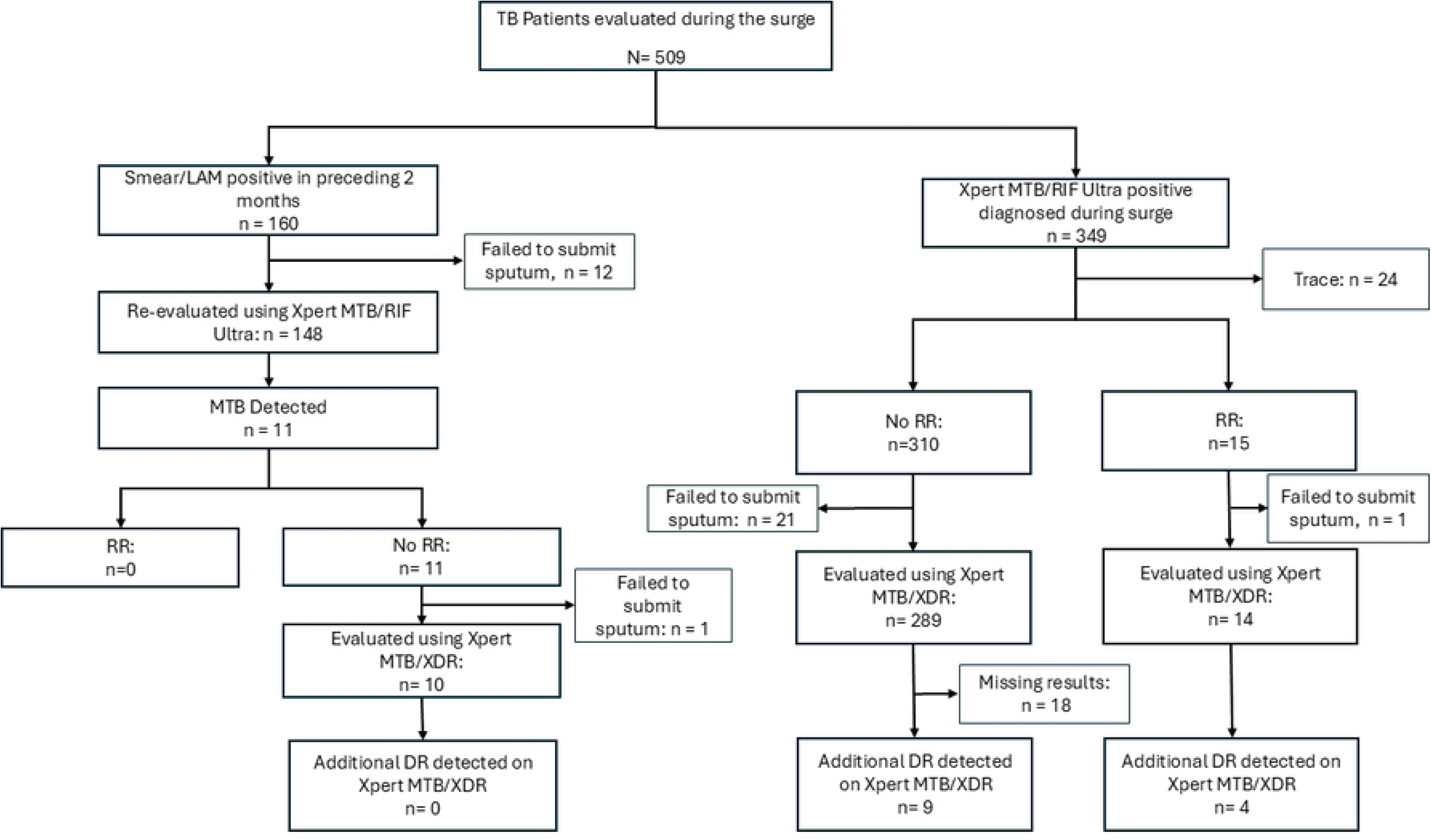
Patient flow chart

### Participant Characteristics

Overall, of the 509 TB patients identified during the surge (median age 38 years, IQR 29–47), the majority were males (70.9%, n=361) and 98.4% (n=498) were aged ≥15 years (Table 1). We identified patients from across all levels of healthcare with the highest representation from health centres (35.8%, n=181) and first-level hospitals (34.1%, n=172). The rest were from second-level hospitals (14.7%, n=74;), and third-level hospitals (n=78; 15.4%). About 60.3% (n=302) of patients were HIV-negative, while only 3.6% (n=18) had unknown HIV status.

**Table 1.** Characteristics of patients included in the study.

| Variable | n (%) |
| --- | --- |
| Age, Median (IQR) | 38 (29-47) |
| Age Group (N - 506)<sup>a</sup> |  |
| 0-4 | 8 (1.6) |
| 5-14 | 11 (2.2) |
| 15+ | 487 (96.2) |
| Sex (N- 506)<sup>a</sup> |  |
| Male | 361(70.9) |
| Female | 148 (29.1) |
| Level of Health Facility (N - 505)<sup>b</sup> |  |
| Health Centre | 181 (35.8) |
| First Level Hospital | 172 (34.1) |
| Second Level Hospital | 74 (14.7) |
| Third Level Hospital | 78 (15.4) |
| HIV Status (n - 501)<sup>c</sup> |  |
| Positive | 180 (35.9) |
| Negative | 302 (63.3) |
| Unknown | 18 (3.6) |

<sup>a</sup>Three (3) missing values; <sup>b</sup>Four (4) missing values; <sup>c</sup>Eight (8) missing values.

### Prevalence of Drug Resistance and Resistance Patterns

While the overall prevalence of DR TB among all bacteriologically confirmed TB patients was 8.5% (75%; n=18 being male.), the prevalence of RR (all forms) was 5.3% (n=15) and that of INH (all forms) was 4.6% (n = 13) (Table 2). Only one patient (0.4%) had combined RR, INH and FQ resistance (Pre-XDR). For mono-resistance among patients with accessible XDR results, 3.5% (n=10) and 3.2% (n=9) were mono-resistant to RIF and INH respectively. Additionally, among all bacteriologically confirmed drug-susceptible TB patients with XDR test results, 1.8% (n=5) had resistance to fluoroquinolones.

**Table 2.** Drug resistance among bacteriologically confirmed TB patients.

| Resistance Pattern | Total | % among all Bact. Confirmed TB with XDR Results (n=282) | % among DR-TB (n=24) |
| --- | --- | --- | --- |
| RR + (INH or FQ) Resistance* | 15 | 5.3 | 62.5 |
| MDR (INH+RR) | 3 | 1.1 | 12.5 |
| INH + RR + FQ | 1 | 0.4 | 4.2 |
| RIF mono-resistance | 10 | 3.5 | 41.7 |
| INH+ (RIF or ETH or FQ) Resistance | 13 | 4.6 | 54.2 |
| INH mono-resistance | 9 | 3.2 | 37.5 |
| INH + ETH | 2 | 0.7 | 8.3 |
| INH + FQ | 1 | 0.4 | 4.2 |
| FQ mono-resistance | 5 | 1.8 | — |

RR – rifampicin resistance, INH – Isoniazid, ETH – Ethionamide, FQ – Fluoroquinolone, \*One (1) RR patient did not submit sputum for XDR testing

Analysis for resistance patterns among DR TB patients (n=24) showed that overall resistance to RIF (all forms) and INH (all forms) was 58.3% and 54.2% respectively. Other resistance patterns observed among DR-TB patients included RIF mono-resistance (n=10, 41.7%), INH mono-resistance (n=9, 37.5%), combined RIF and INH resistance (MDR-TB) (n=3, 12.5%), resistance to INH and ETH (n=2, 8.3%), INH and FQ resistance (n=1, 4.2%), and triple resistance to INH, RIF, and FQ (n=1, 4.2%). In this study, we identified non-RIF resistance in 3.2% (n=9) of all patients with XDR test results (37.5% of DR TB patients).

When stratified by patient category (Table 3), the prevalence of MDR/RR was higher among relapse cases (8.5%, n=4) while INH mono-resistance was higher among new TB patients (3.1%) than among retreatment cases. Among new TB patients (n=223), only 0.4% (n=1) had MDR-TB, while 3.5% (n=8) had RIF mono-resistance, and 3.1% (n=7) had INH mono-resistance. Among TB relapse cases (n=47), 6.4% (n=3) had MDR-TB, 4.3%(n=2) RIF mono-resistance, and 2.1% (n=1) INH mono-resistance. The one patient categorized as treatment after failure had INH mono-resistance while another patient who tested smear positive at two months of treatment remained susceptible to both RIF and INH. All patients treated after loss to follow-up whose results were documented were susceptible to INH and RIF with no other resistance detected.

**Table 3.**
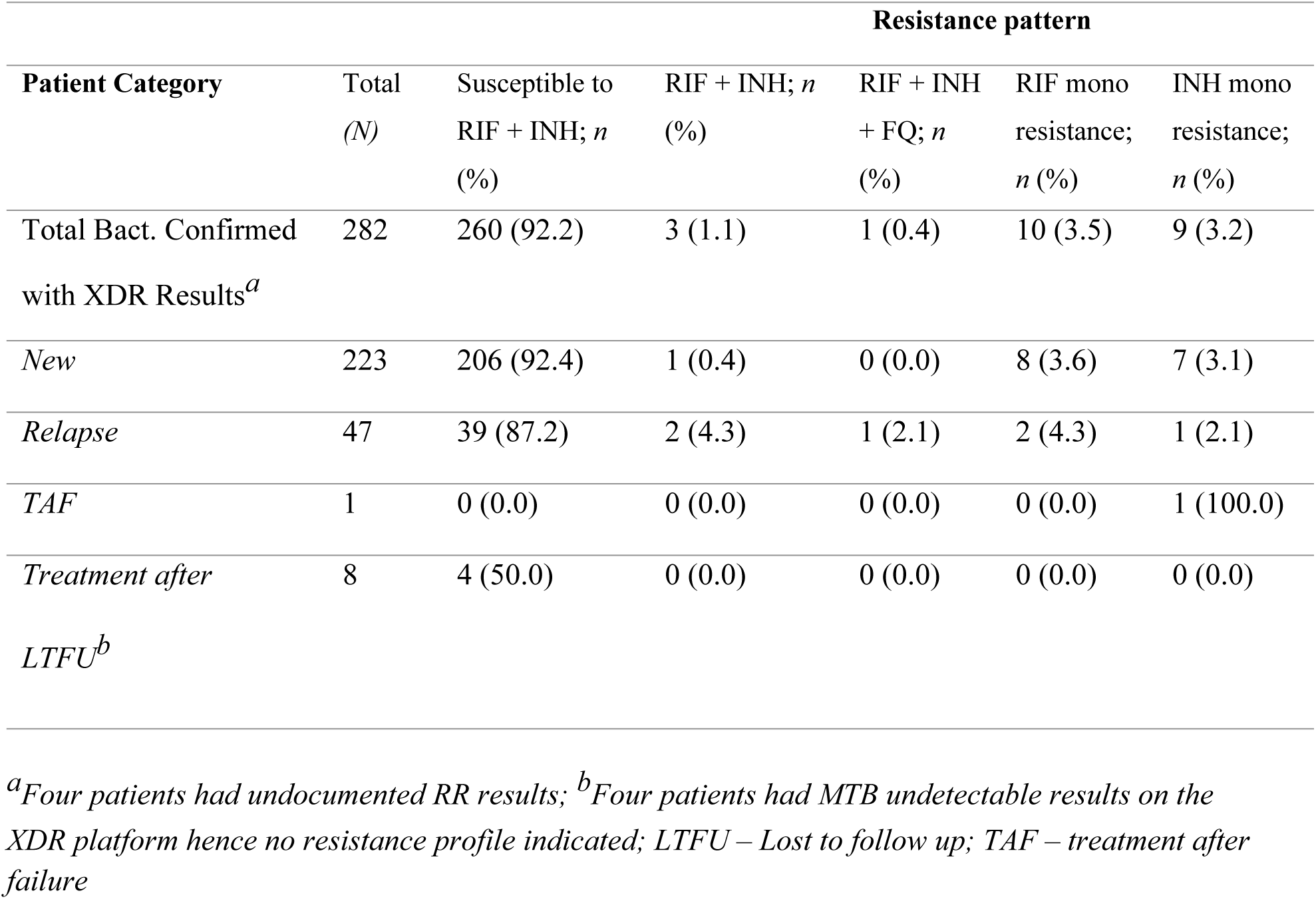
Drug resistance among bacteriologically confirmed TB patients by patient risk category.

## Discussion

In this analysis of programmatic data, we report a DR TB prevalence of 8.5% among bacteriologically confirmed TB patients. Among the DR TB patients, a higher proportion (62.5%) had RR detected on GeneXpert MTB/RIF and over one-third (37.5%) had other forms of resistance detected on GeneXpert MTB/XDR. The prevalence of any INH resistance among patients with accessible XDR test results was 4.6%. One pre-XDR TB patient was diagnosed representing 0.4% of bacteriologically confirmed TB patients with XDR results and 4.2% of the DR TB patients. Of all drug resistance patterns detected, 79.2% had mono-resistance (either RIF or INH). Mono-resistance to either RIF or INH was 5 times more prevalent than MDR-TB in this study.

These findings are similar to the results of Zambia’s most recent DR TB survey where MDR was 8 and 3 times less common than INH and RIF mono-resistance respectively (15). While the national DR-TB survey reported a higher prevalence of INH resistance (3.9%) compared to RIF resistance (1.5%), our study found comparable rates of RIF mono-resistance (3.5%) and INH mono-resistance (3.2%) among TB patients whose XDR test results were accessible. Beside our study finding a lower overall prevalence of drug resistance compared to global estimates of 11.6%, 15.7% and 9.4% for MDR-TB, INH-resistance and RIF Resistance respectively (16), results align with regional and global observations suggesting that mono-resistance remains a significant, and sometimes underestimated, component of the DR-TB burden (16–18).

We also found that over one-third of all DR TB patients detected in this surge had non-RIF resistance patterns: mainly INH with or without FQ and ETH resistance. While the high prevalence of non-rifampicin resistance patterns observed in our study contrasts with the widely held assumption that RIF resistance serves as a reliable proxy for concurrent isoniazid resistance; and thus MDR-TB (19), it aligns with emerging global evidence indicating that a substantial proportion of DR TB cases are RIF-susceptible but resistant to other anti-TB drugs. For example, a multi-country review of 211,753 patients from 156 countries globally found INH mono-resistance prevalence of 8.4% among all TB patients (20). Similarly, a systematic review and meta-analysis with a sample size of 318,430 participants established a pooled INH resistance prevalence of 15.7%; higher than MDR (11.6%) and RIF (9.4%) resistance (16). It is, therefore, possible that TB patients with non-RIF resistance such as INH mono-resistance may often be missed in RIF-centric testing algorithms, hence misclassified and notified as DS TB. This misclassification leads to missed cases of DR TB, thereby undermining DR TB case-finding and prevention efforts. In addition, mis-classified patients are often inappropriately started on INH based first-line regimens. INH resistance requires a tailored treatment approach with possible inclusion of levofloxacin (19), mis-classification results in suboptimal therapy and potentially poor treatment outcomes such as treatment failure, amplification of resistance especially where fluoroquinolone resistance is missed (20–24). Misclassified INH resistance can, therefore, compromise treatment outcomes even when rifampicin is established to be effective (25), delay timely initiation of appropriate treatment, and slow down the interruption of community TB transmission.

Over one-third of bacteriologically confirmed TB patients in this study were identified at primary health care (PHC) facilities, which aligns with existing evidence that nearly half of all TB cases in Zambia are diagnosed at the PHC level (26). That close to 90% of these patients diagnosed at PHC level accessed XDR testing demonstrates the utility of the tool in expanding rapid access to DST at lower levels of care, where access to LPA as well as culture and phenotypic DST (pDST) is often limited low and middle income countries like Zambia (27) despite being the largest contributor to TB notifications. While culture-based phenotypic DST has been useful and remains the gold standard for detecting non-RIF resistance in Zambia and many other countries (28), poor access especially in PHC settings, characterized by longer turnaround time associated with sample courier to centralized laboratories and results transmission back to referring facilities, delays decision-making on the choice of appropriate treatment regimen (29–31). A substantially reduced time to accurate diagnosis achieved through decentralized DST using platforms like Xpert/XDR testing; and prompt initiation of appropriate treatment is, therefore, a principle benefit for both the healthcare workers and TB patients (32).

Although Xpert MTB/XDR assays demonstrate slightly lower sensitivity for detecting ETH and FQ resistance compared to pDST and may fail to capture hetero-resistance patterns (33), they remain valuable for immediate patient management in programmatic settings. Within diagnostic algorithms, Xpert MTB/XDR testing must, therefore, be strategically positioned in near point-of-care settings as a follow-on test for individuals with bacteriologically confirmed TB. This would facilitate quick decision making as results are obtainable within 90 minutes (29). Besides deployment of XDR platforms, utilization must be optimized to get the desired benefit. As done in the surge, systems strengthening: staff capacity building, laboratory logistics availability, sample transportation systems, monitoring/data management systems and ownership by health care workers including those working in the laboratories are required to optimize reflex XDR testing utilization (32). National TB programmes should, therefore, prioritize capacity building of healthcare workers, strengthening of laboratory systems, robust monitoring and data management, and fostering local ownership to optimize the utilization of reflex XDR testing.

This study has several notable limitations and strengths. The short implementation period limited the number of DR TB cases detected, constraining precision in estimating less frequent resistance categories and reducing the scope of analyses that could be conducted. Its focus on urban and peri-urban facilities may not fully capture rural diagnostic realities, while reliance on routine programmatic data could have introduced reporting gaps. Additionally, confirmatory phenotypic DST was not performed. Despite these limitations, the study provides timely, operationally relevant evidence supporting national scale-up of Xpert MTB/XDR testing to enhance rapid detection and management of DR tuberculosis beyond RIF resistance. It represents one of the first programmatic evaluations of the Xpert MTB/XDR assay in Zambia and the region, conducted under real-world MoH conditions across 39 high-burden facilities in five provinces, making the findings both programmatically relevant and generalizable. Integration of digital monitoring tools, logistics support, and staff capacity building further strengthened implementation quality to consider when integrating reflex Xpert MTB/XDR testing into routine diagnostic algorithms.

## Conclusion

The Xpert MTB/XDR assay proved to be a valuable addition to Zambia’s TB diagnostic tools during this nationwide MDR-TB surge with high utility at primary health care level facilities. The ability to detect non-RIF resistance rapidly is essential for accelerating early detection of DR TB, finding missed cases, achieving universal access to DST, optimizing treatment regimens, and improving TB elimination efforts at lower levels of care. We recommend a national scale-up of reflex XDR testing, integrated with robust supply chain management for laboratory commodities, health care workers training, and real-time data use, to advance the fight against DR-TB in Zambia and similar settings.

## Data Availability

All data will be submitted as part of this manuscript

## Declarations

### Availability of Data and materials

All data will be submitted as part of this manuscript

## Funding

There was no specific funding for this project.

### Authors Contributions

Data extraction was done by Innocent Mwaba

Data cleaning and analysis was done by Minyoi Mubita Maimbolwa and Phallon Mwaba The original draft of the manuscript by Phallon Mwaba

Critical review of the manuscript was done by Chitalu Chanda, Minyoi Maimbolwa, Marshal Sikandagwa, Muhammad Mputu, Pauline Kasese-Chanda, Kevin Zimba, Rhehab Chimzizi, Angel Mubanga, Mary Kagujje, and Monde Muyoyeta.

Supervision and validation were done by Mary Kagujje All authors reviewed the manuscript and agreed to publish.

## Acknowledgements

The authors would like to thank the staff at MoH health facilities who were involved in documenting the patients and contacts information in registers from where this data was collected.

